# Association of demographic and occupational factors with SARS-CoV-2 vaccine uptake in a multi-ethnic UK healthcare workforce: a rapid real-world analysis

**DOI:** 10.1101/2021.02.11.21251548

**Authors:** Christopher A. Martin, Colette Marshall, Prashanth Patel, Charles Goss, David R. Jenkins, Claire Ellwood, Linda Barton, Arthur Price, Nigel J. Brunskill, Kamlesh Khunti, Manish Pareek

**Affiliations:** Department of Respiratory Sciences, University of Leicester, UK; Department of Infection and HIV Medicine, University Hospitals of Leicester NHS Trust, Leicester, UK; Deputy Medical Director, University Hospitals of Leicester NHS Trust, Leicester, UK; Department of Chemical Pathology and Metabolic Diseases, University Hospitals of Leicester NHS Trust, Leicester, UK; Department of Cardiovascular Sciences, University of Leicester, UK; Department of Occupational Health, University Hospitals of Leicester NHS Trust, Leicester, UK; Department of Clinical Microbiology, University Hospitals of Leicester NHS Trust, Leicester, UK; Department of Pharmacy, University Hospitals of Leicester NHS Trust, Leicester, UK; Department of Haematology, University Hospitals of Leicester NHS Trust, Leicester, UK; Department of Immunology, University Hospitals of Leicester NHS Trust, Leicester, UK; Department of Nephrology, Leicester General Hospital, Leicester, UK; Diabetes Research Centre, Leicester General Hospital, University of Leicester, Leicester, UK; Leicester Real World Evidence Unit, Diabetes Research Centre, Leicester General Hospital, University of Leicester, Leicester, UK; NIHR Leicester Biomedical Research Centre, Leicester, UK; NIHR Applied Research Collaboration-East Midlands, Leicester, UK

## Abstract

**Background:** Healthcare workers (HCWs) and ethnic minority groups are at increased risk of COVID-19 infection and adverse outcome. Severe acute respiratory syndrome coronavirus-2 (SARS-CoV-2) vaccination is now available for frontline UK HCWs; however, demographic/occupational associations with vaccine uptake in this cohort are unknown. We sought to establish these associations in a large UK hospital workforce.

**Methods:** We conducted cross-sectional surveillance examining vaccine uptake amongst all staff at University Hospitals of Leicester NHS Trust. We examined proportions of vaccinated staff stratified by demographic factors, occupation and previous COVID-19 test results (serology/PCR) and used logistic regression to identify predictors of vaccination status after adjustment for confounders.

**Findings:** We included 19,044 HCWs; 12,278 (64.5%) had received SARS-CoV-2 vaccination. Compared to White HCWs (70.9% vaccinated), a significantly smaller proportion of ethnic minority HCWs were vaccinated (South Asian 58.5%, Black 36.8% p<0.001 for both). After adjustment, factors found to be negatively associated with vaccine uptake were; younger age, female sex, increasing deprivation and belonging to any non-White ethnic group (Black: aOR0.30, 95%CI 0.26–0.34, South Asian:0.67, 0.62–0.72). Those that had previously had confirmed COVID-19 (by PCR) were less likely to be vaccinated than those who had tested negative.

**Interpretation:** Ethnic minority HCWs and those from more deprived areas as well as younger, female staff are less likely to take up SARS-CoV-2 vaccination. These findings have major implications for the delivery of SARS-CoV-2 vaccination programmes in HCWs and the wider population and should inform the national vaccination programme to prevent the disparities of the pandemic from widening.

**Funding:** NIHR, UKRI/MRC

## Introduction

COVID-19, the disease caused by infection with severe acute respiratory syndrome coronavirus-2 (SARS-CoV-2) has spread to become a global pandemic causing significant morbidity and mortality in many countries. As of February 2021, total worldwide COVID-19 cases are estimated to be over 100 million and deaths related to COVID-19 number over 2.1million.^1^ In recent months, thanks to an unprecedented global research effort, a number of vaccines against SARS-CoV-2 have been developed and approved^2,3^ and it is hoped that mass vaccination programmes will aid in slowing transmission of the virus as well as reducing hospitalisation and death from COVID-19.

As the pandemic has progressed, it has become clearer that certain factors may increase the risk of acquiring SARS-CoV-2 infection, including age, obesity and presence of particular comorbidities (e.g. diabetes and cardiovascular disease), occupation and household size.^4–6^ Amongst these ‘high-risk’ groups are healthcare workers (HCWs)^6,7^ in whom an increased risk of hospitalisation with COVID-19 has also been demonstrated.^8^ Within a HCW population, it has been shown that risk of SARS-CoV-2 infection, differs by occupational role and is highest in ‘front-door’ and patient facing specialities,^8,9^ implying that at least some of the increased risk faced by HCWs is mediated through occupational exposure to SARS-CoV-2. In recognition of this risk the UK Joint Committee on Vaccination and Immunisation (JCVI) listed frontline HCWs as a priority group for receiving vaccination against SARS-CoV-2.^10^

The COVID-19 pandemic has also disproportionately affected those from ethnic minority groups, with previous work demonstrating an increased risk of SARS-CoV-2 infection and adverse outcome relative to White individuals.^5,11,12^ Furthermore, HCWs of minority ethnicity have been shown to be at higher risk of infection than their White colleagues.^9,13^

In light of the increased risk of COVID-19 infection and adverse outcome faced by ethnic minority HCWs, concerns have been raised regarding uptake of the SARS-CoV-2 vaccine in this group both in the UK and in US^14,15^. These concerns are founded upon previous work conducted in the general population, which has demonstrated reduced vaccine uptake by ethnic minority individuals^14^ as well as recent survey studies investigating intentions to receive vaccination against COVID-19 which have demonstrated an increased likelihood of vaccine hesitancy in ethnic minority groups.^15,16^

Previous studies in HCW populations have found that occupational factors may contribute to influenza vaccine uptake with medical staff most likely, and healthcare students least likely, to be vaccinated.^17^ However, whether such factors will also be important in predicting COVID-19 vaccine uptake is not known. SARS-CoV-2 vaccination in a HCW cohort is important not only for protection of the individual but, given that a significant proportion of COVID-19 inpatients acquire their infection in hospital and that this has been attributed to HCW to patient transmission, may also prove to be important for reducing nosocomial transmission of COVID-19.^18,19^ It is also currently unknown whether a history of COVID-19 (as defined by positive serology or polymerase chain reaction [PCR] testing) impacts upon vaccine uptake in HCWs.

We, therefore, sought to address this uncertainty by conducting rapid surveillance in a multi-ethnic healthcare workforce cohort at a large UK centre to determine the effects of demographic factors (including ethnicity), occupational factors and previous COVID-19 on SARS-CoV-2 vaccine uptake.

## Methods

### Study design and study centre

This cross-sectional surveillance was conducted at University Hospitals of Leicester (UHL) NHS Trust, one of the largest acute hospital trusts in the UK, where 36% of staff are from minority ethnic backgrounds.^20^ UHL is the only acute hospital trust serving the population of Leicester, Leicestershire and Rutland (approximately 1 million residents) and cares for the vast majority of hospital attenders with COVID-19 from these areas. Leicester has seen comparatively high rates of SARS-CoV-2 transmission across the course of the pandemic compared to other areas of the UK and has been subject to ‘lockdown’ measures since June 2020 ^21,22^

### Staff vaccination programme

UHL began vaccinating staff against SARS-CoV-2 on 12^th^ December 2020, initially using the BNT162b2 mRNA (Pfizer-BioNTech)^2^ and subsequently also with the ChAdOx1 nCoV-19 (Oxford-AstraZeneca)^3^ COVID-19 vaccines. Three vaccination ‘hubs’ (one at each of the three main hospital sites that make up UHL) were established on the 12^th^ December 2020, 8^th^ January 2021 and 15^th^ of January 2021. Following a short period during which vaccination was limited to those determined to be at highest risk of severe COVID-19, vaccinations were made available to all staff on an ongoing basis. All staff at UHL have received an email inviting them to attend for vaccination and have also received regular reminders to book vaccination appointments via trust-wide electronic and verbal cascaded communications. Line managers were instructed to publicise vaccination, particularly in areas where there is a known low rate of internet or smart phone usage by staff.

### Study population

We included all staff identified in the Electronic Staff Record (ESR) which encompasses all permanent, part-time and bank workers employed by UHL on 3^rd^ February 2021.

### Data collection

We extracted information concerning age (categorised into groups of ≤30, 31 – 40, 41 – 50, 51 – 60 and ≥61 years old); sex; self-reported ethnicity (categorised into White, South Asian, Black and Other for the main analysis and into 15 ethnicity categories for a more granular subanalysis – Supplementary Table 1); occupational role (categorised into 7 categories – Supplementary Table 2) and residential postcode from the ESR. We used residential postcode to obtain the Index of Multiple Deprivation (IMD) quintile using an online tool provided by the UK government. IMD is the official measure of relative deprivation for small areas of the UK. ^23^

We combined these data with information from vaccination centre records to determine SARS-CoV-2 vaccination status and date of vaccination as well as with occupational health records to determine the number, date and result of any SARS-CoV-2 polymerase chain reaction (PCR) or anti-SARS-CoV-2 serology tests as well as the reason given for any recorded COVID-19 related absences from work since the start of the pandemic.

### Data analysis

All variables were categorical and were summarised as count and percentage. Demographic and occupational characteristics were compared using chi-squared test. Number and percentage of staff vaccinated in each week from the start of the vaccination programme to the date of data extraction were plotted. We used logistic regression to evaluate unadjusted and adjusted associations of demographic and occupational factors as well as previous test results for COVID-19 (PCR or serology) with SARS-CoV-2 vaccine uptake.

Multiple imputation was used to replace missing data in all logistic regression models, the multiple imputation model included all variables bar those being imputed. Rubin’s Rules were used to combine the parameter estimates and standard errors from 10 imputations into a single set of results. ^24^

All analyses were conducted using Stata (StataCorp. 2017. Stata Statistical Software: Release 16.1 College Station, TX: StataCorp LLC). P-values <0.05 were considered statistically significant. Figures were prepared in Excel (Microsoft 2021).

### Ethics

We consulted the NHS Health Research Authority decision aid to ascertain whether ethical approval was required. It was deemed that, as this work represents a service evaluation/surveillance which utilises data collected as part of the routine delivery of a clinical service, approval was not required. In addition, we confirmed approval from our Caldicott Guardian to undertake this work as an audit (UHL11113).

## Results

### Demographic and occupational characteristics of the cohort

In total, 19,044 HCWs were included in the final analysis (see table 1). 9088 (47.7%) were under 40 years of age and 14,395 (75.6%) were female. 11,485 (60.3%) were White, 4863 (25.5%) were of South Asian ethnicity and 1357 (7.1%) of Black ethnicity. Number of vaccinations per week peaked in the week 11^th^ Jan – 17^th^ Jan 2021 and have been in decline since (Figure 1).

**Table 1.** Description of the cohort by vaccination status.

| **Variable** | **Total**<br>**n=19,044** | **Unvaccinated**<br>**n=6,766**<br>**(35.5%)** | **Vaccinated**<br>**n=12,278**<br>**(64.5%)** |
| --- | --- | --- | --- |
| **Age (years)** |  |  |  |
| ≤30 | 4432 (23.3%) | 2142 (31.7%) | 2290 (18.7%) |
| 31 – 40 | 4656 (24.5%) | 1975 (29.2%) | 2681 (21.8%) |
| 41 – 50 | 4312 (22.6%) | 1275 (18.8%) | 3037 (24.7%) |
| 51 – 60 | 4101 (21.5%) | 975 (14.4%) | 3126 (25.5%) |
| ≥61 | 1543 (8.1%) | 399 (5.9%) | 1144 (9.3%) |
| **Sex** |  |  |  |
| Female | 14395 (75.6%) | 5099 (75.4%) | 9296 (75.7%) |
| Male | 4649 (24.4%) | 1667 (24.6%) | 2982 (24.3%) |
| **Ethnicity** |  |  |  |
| White | 11485 (60.3%) | 3338 (49.3%) | 8147 (66.4%) |
| South Asian | 4863 (25.5%) | 2020 (29.9%) | 2843 (23.2%) |
| Black | 1357 (7.1%) | 858 (12.7%) | 499 (4.1%) |
| Other | 1038 (5.4%) | 429 (6.3%) | 609 (5.0%) |
| Not stated | 301 (1.6%) | 121 (1.8%) | 180 (1.5%) |
| **IMD quintile** |  |  |  |
| 5 (least deprived) | 4597 (24.3%) | 1323 (19.6%) | 3274 (26.7%) |
| 4 | 4010 (21.2%) | 1265 (18.7%) | 2745 (22.4%) |
| 3 | 3302 (17.4%) | 1175 (17.4%) | 2127 (17.3%) |
| 2 | 4085 (21.6%) | 1682 (24.8%) | 2403 (19.6%) |
| 1 (most deprived) | 2940 (15.5%) | 1252 (18.5%) | 1688 (13.8%) |
| Missing | 110 (0.6%) | 69 (1.0%) | 41 (0.3%) |
| **Occupation** |  |  |  |
| Doctor | 3001 (15.8%) | 1280 (18.9%) | 1721 (14.0%) |
| Nurse / HCA | 7815 (41.0%) | 2929 (43.3%) | 4886 (39.8%) |
| Allied Health Professional | 1380 (7.3%) | 427 (6.3%) | 953 (7.8%) |
| Admin / executive | 3465 (18.2%) | 928 (13.7%) | 2537 (20.7%) |
| Healthcare Scientist | 871 (4.6%) | 237 (3.5%) | 634 (5.2%) |
| Estates / Facilities | 2306 (12.1%) | 907 (13.4%) | 1399 (11.4%) |
| Other | 206 (1.1%) | 58 (0.9%) | 148 (1.2%) |
| **Previous SARS-CoV-2 serology** |  |  |  |
| Never tested | 7456 (39.2%) | 3656 (54.0%) | 3800 (31.0%) |
| Negative | 10314 (54.2%) | 2732 (40.4%) | 7582 (61.8%) |
| Positive | 1274 (6.7%) | 378 (5.6%) | 896 (7.3%) |
| **Previous SARS-CoV-2 PCR** |  |  |  |
| Never tested | 15136 (79.5%) | 5710 (84.4%) | 9426 (76.8%) |
| Negative | 3072 (16.1%) | 761 (11.3%) | 2311 (18.8%) |
| Positive | 836 (4.4%) | 295 (4.4%) | 541 (4.4%) |
| **Previous COVID-19 work absence** |  |  |  |
| No absence | 12619 (66.3%) | 4749 (70.2%) | 7870 (64.1%) |
| Symptomatic | 3698 (19.4%) | 1221 (18.1%) | 2477 (20.2%) |
| Household / test and trace contact | 2727 (14.3%) | 796 (11.8%) | 1931 (15.7%) |

IMD – Index of Multiple Deprivation

**Figure 1.**
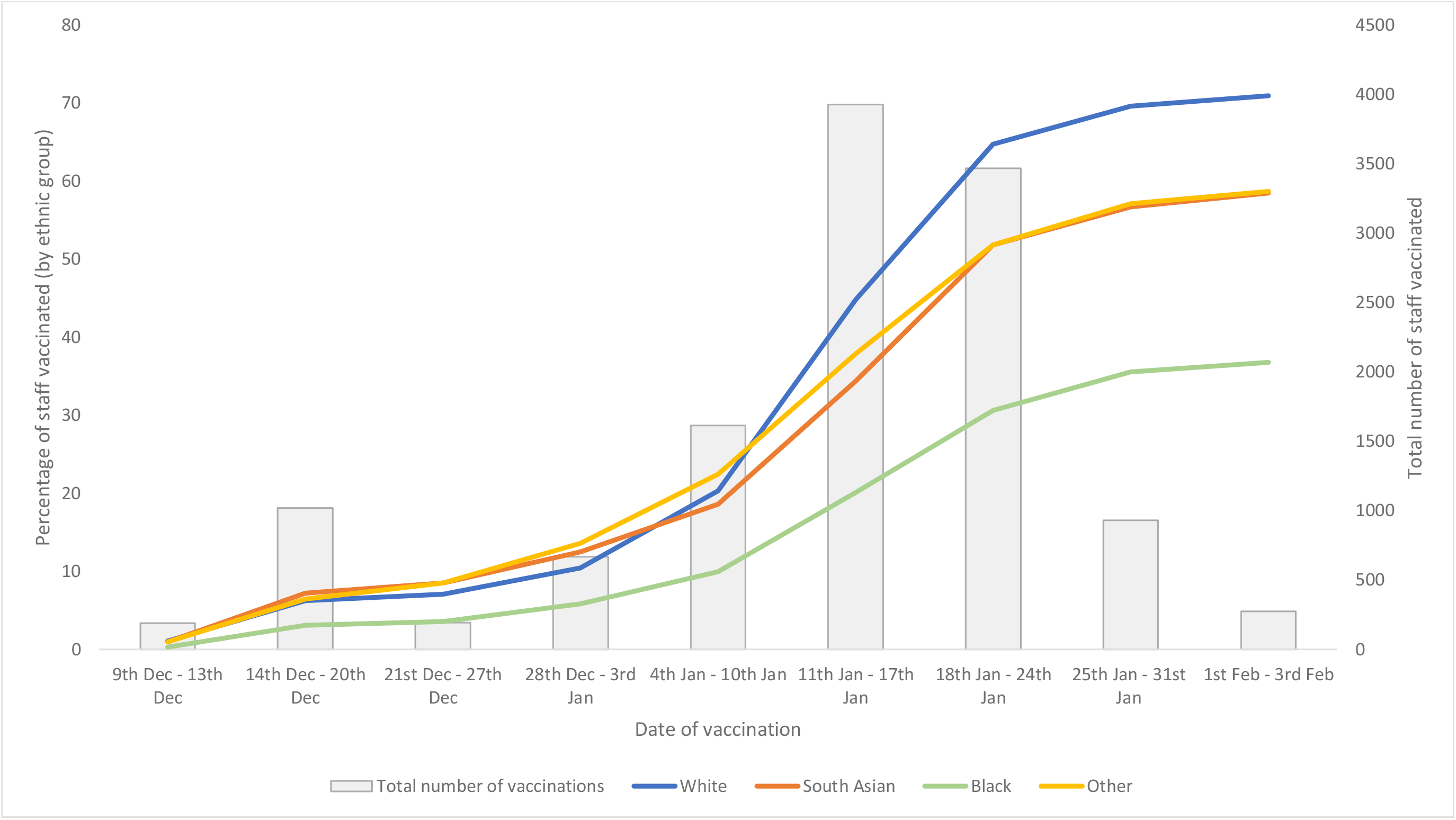
Number and percentage of staff vaccinated over time by ethnic group. Figure shows the number of staff vaccinated (grey bars) and the cumulative percentage of total number of staff of each ethnic group vaccinated (coloured lines) each week since the start of the UHL vaccination programme. It should be noted that the first and last timepoints do not represent a complete week.

### Impact of demographic and occupational factors on SARS-CoV-2 vaccination uptake

Of the 19,044 HCWs in the cohort, 12,278 (64.5%) had received SARS-CoV-2 vaccination. Unvaccinated HCWs were younger than vaccinated HCWs (31.7% of unvaccinated HCWs were under 30 years old compared to 18.7% in the vaccinated cohort [p<0.001]).

Compared to White HCWs (70.9% vaccinated), a significantly lower proportion of ethnic minority HCWs were vaccinated (South Asian 58.5%, Black 36.8% p<0.001 for both; see Table 1 and Figure 1). Within the South Asian cohort, a significantly smaller proportion of Pakistani and Bangladeshi HCWs were vaccinated compared to the Indian cohort (43.2% vs 60.3% and 36.8% vs 60.3% respectively, p<0.001 for both observations). The proportions of vaccinated Black Caribbean and Black African HCWs were similar (39.7% vs 36.2% respectively, p=0.32) – Supplementary Table 3.

The unvaccinated cohort had a greater proportion of HCWs living in areas corresponding to the lower three IMD quintiles (61.4% vs 50.8%, p<0.001 – Table 1).

The occupational groups with the lowest proportions of vaccinated HCWs were doctors (57.4%), estates and facilities staff (60.7%) and nurses and HCAs (62.5%). The occupational group with the highest proportion of vaccinated staff comprised those in administrative and executive roles (73.2%) – see Table 1. In a sensitivity analysis excluding those with locum or bank contracts, vaccination uptake amongst doctors was higher (69.4%) – Supplementary Table 4. A more granular analysis of vaccination uptake in medical staff is shown in Supplementary Table 5. Within medical staff, vaccination uptake is highest amongst consultants (81.5% vaccinated) and foundation year 1 doctors (69.6% vaccinated) and lowest amongst senior house officers (SHOs) / speciality registrars (42.5% vaccinated), medical support staff (31.9% vaccinated) and the small number of general practitioners (9.5% vaccinated).

### Association of prior SARS-CoV-2 infection with vaccination uptake

11,588/19,044 (60.8%) staff had previously been tested for anti-SARS-CoV-2 IgG, 1274 (11.0%) of these were positive. 3908/19,044 (20.5%) had previously undergone nasopharyngeal PCR testing for SARS-CoV-2, 836 (21.4%) of these were positive.

When only staff who had undergone serological testing were included, staff with detectable antibody formed a greater proportion of the unvaccinated cohort compared to the vaccinated cohort (12.2% vs 10.6%, p=0.02). Proportions of those with a previous positive SARS-CoV-2 PCR test in the vaccinated and unvaccinated cohorts were the same (4.4% for both).

### Factors associated with SARS-CoV-2 vaccine uptake

Table 2 shows univariable and multivariable (adjusted for age, sex, ethnicity, IMD, occupation, previous SARS-CoV-2 testing and work absences) logistic regression models for factors associated with vaccination against SARS-CoV-2.

**Table 2.**
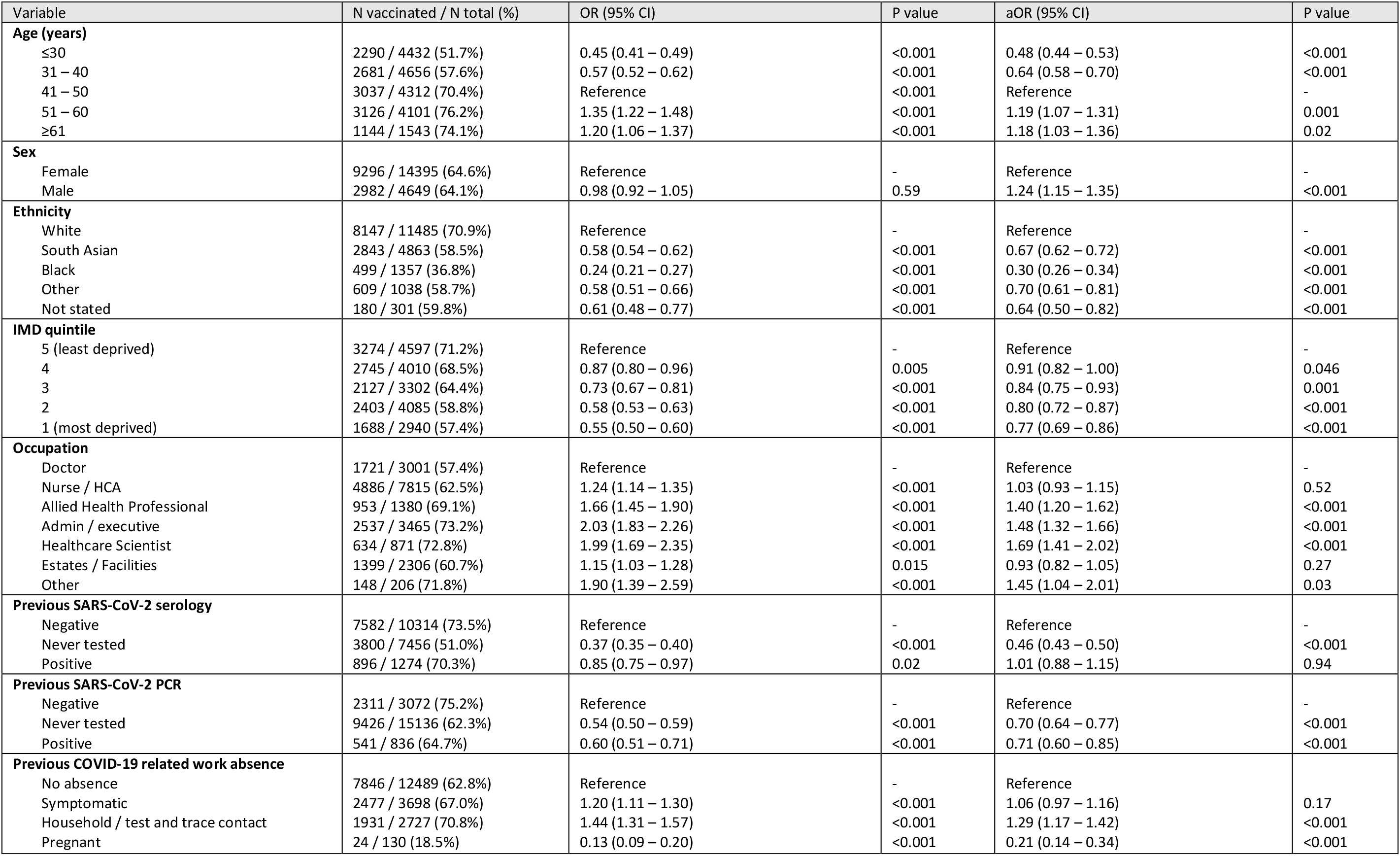
Univariable and multivariable analysis of factors associated with SARS-CoV-2 vaccine uptake.

After adjustment, factors associated with uptake of vaccination included increasing age (age group ≤30 years: aOR 0.48 95%CI 0.44 – 0.53; age group 51 – 60 years: 1.19, 1.07 – 1.31 compared to age 41 - 50) and male sex (aOR 1.24 95%CI 1.15 – 1.35).

HCWs from ethnic minority backgrounds were significantly less likely than their White colleagues to be vaccinated, an effect most marked in those of Black ethnicity (Black: aOR 0.30, 95%CI 0.26 – 0.34, South Asian: aOR 0.67, 95%CI 0.62 – 0.72). We found that vaccination uptake decreased with increasing deprivation (decrease in IMD quintile; test for trend p<0.001).

In comparison to doctors, allied health professionals (aOR 1.40, 95%CI 1.20 – 1.62), administrative and executive staff (1.48, 1.32 – 1.66) and healthcare scientists (1.69, 1.41 – 2.02) were all around 1.5 times more likely to be vaccinated. However, in a sensitivity analysis excluding those with locum/bank contracts (Supplementary Table 6) doctors were not less likely than other groups to be vaccinated and were, in fact, more likely than nurses/HCAs and estates/facilities staff to take up vaccination. Other significant findings remained unchanged.

Staff who had never undergone serology or PCR testing for SARS-CoV-2 were significantly less likely to have been vaccinated than those who had tested negative (serology: aOR 0.70, 95%CI 0.64 - 0.77 and PCR: 0.70, 0.64 – 0.77). Those staff with a history of a previous positive SARS-CoV-2 PCR result were significantly less likely to be vaccinated than those with only negative results (aOR 0.71, 95% CI 0.60 – 0.85). To ensure that this effect was not simply due to these individuals not accessing vaccination due to recent COVID-19 infection we conducted a sensitivity analysis excluding individuals who tested positive for SARS-CoV-2 by PCR after a date 28 days prior to the vaccine programme starting (n=289) from the multivariable model. The significant findings remained unchanged.

## Discussion

In this observational analysis in one of the largest and most ethnically diverse populations of HCWs in the UK, we found SARS-CoV-2 vaccine uptake to be significantly lower in those of younger age groups, female sex and those living in more deprived areas. We also found that ethnic minority HCWs are significantly less likely to take up vaccination than those of White ethnicity and that this difference is particularly marked for Black HCWs and certain South Asian HCW groups.

To our knowledge, we are the first to provide real-life data demonstrating that SARS-CoV-2 vaccine uptake is reduced in ethnic minority HCWs. This adds significant weight to emerging data in the general population which also suggests reduced uptake in ethnic minority groups. Our findings closely align with this population level data as we also demonstrate that those of Black ethnicity were least likely to take up vaccination and that, amongst South Asian ethnic groups, those of Pakistani and Bangladeshi ethnicity were less likely to take up SARS-CoV-2 vaccination than those of Indian ethnicity^25^. Evidence on the barriers to COVID-19 vaccination in ethnic minority groups is limited^14^. However, when vaccine uptake is considered more broadly, factors such as a lack of trust in the government or in healthcare systems (e.g. due to unethical and non-ethnically heterogenous research practices in vaccine studies or structural and institutional racism), a lower perception of the risk of COVID-19 or a higher perception of the risk of side effects from vaccination and other sociodemographic factors interrelated to ethnicity (educational level, socioeconomic status and religion) have all been suggested as barriers to vaccine uptake.^14,26–28^ Regardless of the underlying mechanisms, these findings give significant cause for concern, as ethnic minority groups (especially those working in healthcare) are at higher risk of infection with SARS-CoV-2 and adverse outcome from COVID-19, yet are not taking up this critical preventative intervention^3,5,9,11^. Furthermore, HCWs are an important source of health information for ethnic minority communities^14^ and so our findings may also have implications for vaccine uptake in the population at large.

Alongside ethnicity, we also found deprivation to be associated with SARS-CoV-2 vaccination uptake, with those living in the most deprived areas being most likely to be unvaccinated. Deprivation has previously been shown to be associated with lower vaccine uptake in the general UK population^29^ and this may be mediated through many of the same mechanisms discussed in relation to ethnicity above.

Younger healthcare workers were also less likely to be vaccinated than their older colleagues; a likely explanation is a reduced perception of personal risk of adverse outcome from COVID-19. However, alongside the obvious greater risk of transmitting infection to more vulnerable individuals, long-term sequelae of COVID-19 (termed ‘long-COVID’) which may cause significant morbidity, has been demonstrated to be prevalent even in a young ‘low-risk’ population,^30^ suggesting that this cohort may still derive significant personal benefit from SARS-CoV-2 vaccination. A further explanation for this finding is that the vaccination programme was initially targeted at those with risk factors for severe COVID-19 (including those advanced in age) and thus older staff may have had more time and opportunity to be vaccinated compared to their younger colleagues.

We found that doctors were significantly less likely to take up SARS-CoV-2 vaccination than other staff groups (including allied health professionals). However, these findings should be interpreted with caution, as exclusion of individuals with locum/bank contracts resulted in higher uptake amongst doctors. It is possible, therefore, that locum doctors are not taking up vaccination through UHL. This may be due to limited access to trust communications or due to taking up offers of vaccination elsewhere. Further work is required to determine vaccine uptake in doctors, and if our findings are repeated in other studies then determination of barriers to SARS-CoV-2 vaccination in doctors should be a focus of future studies. Estates and facilities staff also had lower levels of vaccine uptake than many other groups, support staff have been found to have low levels of vaccine uptake previously^28^ and possible explanations for this observation in our cohort include limited access to the email communications regarding vaccination, as well as factors interrelated to occupational role such as educational level, deprivation and ethnicity.

We also investigated the relationship of previous COVID-19 infection to SARS-CoV-2 vaccine uptake in a population of HCWs. We found that those who were never tested for evidence of current/previous SARS-CoV-2 infection by swab or serology were more likely to be unvaccinated. This is unsurprising given that many of the barriers to vaccination (e.g. mistrust of the healthcare system, ‘needle phobia’ and low perception of personal risk from COVID-19) may also influence decisions about testing for evidence of current/previous SARS-CoV-2. Those with a history of SARS-CoV-2 PCR positivity were also less likely to take up vaccination. Some of this effect could be mediated by those who were isolating due to a positive swab having no access to vaccination as well as advice from the trust that those with a positive swab in the last 28 days should avoid vaccination. However, exclusion of individuals testing positive after a date 28 days prior to the start of the vaccination programme did not change the result implying the influence of other factors. It is possible that some of those who have had confirmed COVID-19 would be less likely to take up vaccination, believing themselves to have acquired sufficient immunological protection to SARS-CoV-2. This is likely to be true in the short term; however, risk of infection may increase with time since infection, given evidence concerning waning humoral immunity to SARS-CoV-2 and the short lived immunity after infection with other coronaviruses.^32,33^ therefore this group may represent an important group to target in subsequent SARS-CoV-2 vaccination drives.

This study has limitations. Although the population is large, data are from a single centre affecting their generalisability. We only have vaccination data on those who were vaccinated through UHL. HCWs who obtained vaccination through primary care will be coded as unvaccinated in our analysis, although we expect these numbers to be small given that few other vaccination centres were in operation prior to establishing vaccination hubs at UHL. We cannot predict if HCWs who are currently unvaccinated will take up vaccination in the future, however the numbers of staff taking up the vaccine over time are falling, implying that most who will accept vaccination have already done so. SARS-CoV-2 PCR testing has been available at other non-UHL centres and PCR results from HCWs accessing testing via these centres were not available within UHL records, however given the convenience and availability of PCR testing within UHL it is likely that the vast majority of staff would have accessed testing via this route. There are other factors which may influence vaccine uptake, (e.g. past medical history, educational level) on which we do not have data as we felt this was beyond the scope of an audit, and therefore cannot adjust for these in our analysis. Despite these limitations, our work has many novel findings which will be of direct relevance to policy makers involved in designing SARS-CoV-2 vaccination programmes.

In summary, we have found that in a population of UK HCWs, those from ethnic minority groups and from more deprived areas, as well as younger females and those from particular occupational groups are less likely to take up SARS-CoV-2 vaccination. These findings have major implications for the effective ongoing delivery of SARS-CoV-2 vaccination programmes, both in HCWs and in the wider population, and should be acted upon urgently to prevent the disparities caused by the COVID-19 pandemic from being allowed to widen further.

## Data Availability

The patient cohort was extracted under Caldicott Guardian approval for a specific purpose and as part of our undertaking with them we are not to further routinely share this data. The data is held in an institutional repository and interested parties, with appropriate approvals, can apply for data access through the Corresponding author. Reasonable requests will be assessed on a case-by-case basis in discussion with the Caldicott Guardian.

## Author Contributions

CM, PP and CG collected the data. CAM analysed the data with input from MP. CAM drafted the manuscript with input from MP. All authors discussed the results and contributed to the final manuscript.

## Acknowledgements

Tony Doherty and Michael Dobson assisted with data collection. Matt Archer was involved in establishing the UHL vaccination programme.

