## Supplementary information for "Association of demographic and occupational factors with SARS-CoV-2 vaccine uptake in a multi-ethnic UK healthcare workforce: a rapid real-world analysis"

**Running title**

SARS-CoV-2 vaccination in healthcare workers

**Supplementary table 1. Categorisation of ethnicity**

| **ETHNICITY (MANUSCRIPT)** | **ETHNICITY (ELECTRONIC STAFF RECORD)** |
| --- | --- |
| White | A White – British, B White – Irish, C White - Any other White background |
| South Asian | "H Asian or Asian British - Indian","J Asian or Asian British - Pakistani", "L Asian or Asian British - Bangladeshi", "K Asian or Asian British - Bangladeshi","L Asian or Asian British - Any other Asian background" |
| Black | "M Black or Black British - Caribbean", "N Black or Black British - African", "P Black or Black British - Any other Black background" |
| Other | "D Mixed - White & Black Caribbean", "E Mixed - White & Black African", "F Mixed - White & Asian", "G Mixed - Any other mixed background”, "R Chinese" |
| Not Stated | “Z Not Stated” |

**Supplementary table 2. Categorisation of occupational role**

| **OCCUPATIONAL ROLE (MANUSCRIPT)** | **OCCUPATIONAL ROLE (HEALTH RECORD)** |
| --- | --- |
| Doctors | "Consultant", "Foundation Year 1" ,"Core Trainee - Old Age", "Foundation Year 2", "Senior House Officer" "Specialist Registrar", "Specialty Doctor", "Specialty Registrar", Staff Grade", "Trust Grade Doctor - Career Grade level", "Trust Grade Doctor - Foundation Level", "Trust Grade Doctor - SHO Level"  "Trust Grade Doctor - Specialty Registrar", "Physician Associate", "Advanced Practitioner, "Associate Specialist (Closed to new entrants)", "Emergency Care Practitioner", "General Medical Practitioner", "Claims Doctor", "STR", "Specialist Registrar", "Trust Grade Doctor - Specialist Registrar", "Salaried General Practitioner", "Medical Student", “Hospital Practitioner”, “Clinical Assistant” |
| Nurses / HCAs | "Nurse Consultant", "Nurse Manager", "Nurse Qualified", "Nurse Qualified Hospital L1", "Nursery Nurse", "Nursing Associate", "Sister/Charge Nurse", "Staff Nurse", "Unqualified Nurse", "Health Care Assistant", "Healthcare Assistant", "Bank Midwife", "Midwife", "Midwife - Manager", "Midwife - Specialist Practitioner", "Modern Matron", "Enrolled Nurse", "Trainee Nursing Associate", "Trainee Practitioner", "Deputy Sister/Charge Nurse", "Assistant Practitioner Nursing", "Assistant/Associate Practitioner", "Assistant/Associate Practitioner Nursing", "Health Care Support Worker", "Director of Nursing" "Ward Sister/Charge Nurse" |
| Allied Health Professionals | "Occupational Therapist", "Occupational Therapist Manager", "Occupational Therapy Specialist Practitioner", "Physiotherapist", "Physiotherapist Manager", "Physiotherapist Specialist Practitioner", "Operating Department Practitioner", "Phlebotomist", "Pharmacist", "Pre-reg Pharmacist", “Student Technician – Pharmacy” "Radiographer - Diagnostic", "Radiographer - Diagnostic, Consultant", "Radiographer - Diagnostic, Manager", "Radiographer - Diagnostic, Specialist Practitioner", "Radiographer - Therapeutic", “Student Technician –Trauma and Orthopaedic surgery” |
| Administrative/Executive | "Secretary", "Manager", "Accountant", "Medical Secretary", "Apprentice", "Officer", "Clerical Worker", "Board Level Director", “Chair”, "Librarian", "Finance Director", "Other Executive Director", "Receptionist", "18 Week Pathway Co-ordinator", "Adviser", "Senior Manager", "Personal Assistant", "Analyst", "Systems Manager" |
| Healthcare Scientists | "Healthcare Science Assistant", "Healthcare Science Associate", "Healthcare Science Practitioner", "Healthcare Scientist", "Researcher", "Consultant Healthcare Scientist", "Biomedical Scientist”, "Trainee Healthcare Science Associate", "Trainee Healthcare Science Practitioner ", "Trainee Healthcare Scientist", "Specialist Healthcare Scientist", "Specialist Healthcare Science Practitioner", “Student Technician – Clinical Cytogenetics” |
| Estates | "Assistant", "Building Craftsperson", "Carpenter", "Cleaner", "Cook", "Driver", "Electrician", "Engineer", "Gardener/Groundsperson", "Housekeeper", "Maintenance Craftsperson", "Porter", "Supervisor", "Technician", "Ward Housekeeper" |
| Other | "Chaplain", "Helper/Assistant", "Interpreter", "Network Engineer", "Child and Adolescent Psychiatry""Forensic Psychiatry", "General Practice", "General Psychiatry", "Medical Psychotherapy", "Old Age Psychiatry", "Occupational Health", "Pain Management", "Psychiatry", "Psychiatry of Learning Disability", "Rehabilitation Psychiatry", "Sport and Exercise Medicine", "Substance Abuse", |

**Supplementary Table 3. Vaccination status by ethnicity subcategory**

| **Ethnicity** | **Unvaccinated, n(%)** | **Vaccinated, n(%)** |
| --- | --- | --- |
| White British / Irish | 2794 (27.8%) | 7262 (72.2%) |
| Other White | 544 (38.1%) | 885 (61.9%) |
| Indian | 1477 (39.8%) | 2239 (60.3%) |
| Pakistani | 249 (56.9%) | 189 (43.2%) |
| Bangladeshi | 74 (63.3%) | 43 (36.8%) |
| Other South Asian | 220 (37.2%) | 372 (62.8%) |
| Black Caribbean | 137 (60.4%) | 90 (39.7%) |
| Black African | 641 (63.8%) | 363 (36.2%) |
| Other Black | 80 (63.5%) | 46 (36.5%) |
| Mixed White / Black | 96 (47.8%) | 105 (52.2%) |
| Mixed White / Asian | 39 (42.9%) | 52 (57.1%) |
| Other Mixed | 45 (43.3%) | 59 (56.7%) |
| Chinese | 61 (40.4%) | 90 (59.6%) |
| Other | 188 (38.3%) | 303 (61.7%) |
| Not stated | 121 (40.2%) | 180 (59.8%) |

| **Variable** | **Total**  n=16,433 | **Unvaccinated**  n=4917  (29.9%) | **Vaccinated**  n=11,516  (70.1%) |
| --- | --- | --- | --- |
| **Age (years)**  ≤30  31 – 40  41 – 50  51 – 60  ≥61 | 3490 (21.2%)  4014 (24.4%)  3873 (23.6%)  3791 (23.1%)  1265 (7.7%) | 1430 (29.1%)  1477 (30.0%)  970 (19.7%)  802 (16.3%)  238 (4.8%) | 2060 (17.9%)  2537 (22.0%)  2903 (25.2%)  2989 (26.0%)  1027 (8.9%) |
| **Sex**  Female  Male | 12625 (76.8%)  3808 (23.2%) | 3866 (78.6%)  1051 (21.4%) | 8759 (76.1%)  2757 (23.9%) |
| **Ethnicity**  White  South Asian  Black  Other  Not stated | 10097 (61.4%)  4203 (25.6%)  1034 (6.3%)  870 (5.3%)  229 (1.4%) | 2426 (49.3%)  1536 (31.2%)  583 (11.9%)  297 (6.0%)  75 (1.5%) | 7671 (66.6%)  2667 (23.2%)  451 (3.9%)  573 (5.0%)  154 (1.3%) |
| **IMD quintile**  5 (least deprived)  4  3  2  1 (most deprived)  Missing | 3998 (24.3%)  3486 (21.2%)  2850 (17.3%)  3526 (21.5%)  2516 (15.3%)  57 (0.4%) | 933 (19.0%)  906 (18.4%)  875 (17.8%)  1254 (25.5%)  931 (18.9%)  18 (0.4%) | 3065 (26.6%)  2580 (22.4%)  1975 (17.2%)  2272 (19.7%)  1585 (13.8%)  39 (0.3%) |
| **Occupation**  Doctor  Nurse / HCA  Allied Health Professional  Admin / executive  Healthcare Scientist  Estates / Facilities  Other | 2299 (14.0%)  6669 (40.6%)  1281 (7.8%)  3209 (19.5%)  806 (4.9%)  1978 (12.0%)  191 (1.2%) | 704 (14.3%)  2157 (43.9%)  364 (7.4%)  768 (15.6%)  198 (4.0%)  678 (13.8%)  48 (1.0%) | 1595 (13.9%)  4512 (39.2%)  917 (8.0%)  2441 (21.2%)  608 (5.3%)  1300 (11.3%)  143 (1.2%) |
| **Previous SARS-CoV-2 serology**  Never tested  Negative  Positive | 5611 (34.1%)  9669 (58.8%)  1153 (7.0%) | 2250 (45.8%)  2363 (48.1%)  304 (6.2%) | 3361 (29.2%)  7306 (63.4%)  849 (7.4%) |
| **Previous SARS-CoV-2 PCR**  Never tested  Negative  Positive | 12,765 (77.7%)  2, 886(17.6%)  782 (4.8%) | 4018 (81.7%)  638 (13.0%)  261 (5.3%) | 8747 (76.0%)  2248 (19.5%)  521 (4.5%) |
| **Previous COVID-19 work absence**  No absence  Symptomatic  Household / test and trace contact  Pregnant | 9878 (60.1%)  3698 (22.5%)  2727 (16.6%)  130 (0.8%) | 2794 (56.8%)  1221 (24.8%)  796 (16.2%)  106 (2.2%) | 7084 (61.5%)  2477 (21.5%)  1931 (16.8%)  24 (0.2%) |

**Supplementary Table 4. Description of cohort excluding locum or bank staff**

**Supplementary Table 5. Vaccine uptake in medical staff**

| **Grade of medical staff** | **Total cohort** | | **Locum and bank workers excluded** | |
| --- | --- | --- | --- | --- |
|  | **Unvaccinated** | **Vaccinated** | **Unvaccinated** | **Vaccinated** |
| **FY1** | 35 (30.4%) | 80 (69.6%) | 34 (29.8%) | 80 (70.2%) |
| **FY2** | 50 (37.9%) | 82 (62.1%) | 50 (37.9%) | 82 (62.1%) |
| **SHO/SpR** | 689 (57.5%) | 509 (42.5%) | 287 (39.5%) | 439 (60.5%) |
| **Consultant** | 164 (18.6%) | 720 (81.5%) | 154 (17.7%) | 718 (82.3%) |
| **Trust grade** | 168 (41.3%) | 239 (58.7%) | 138 (37.1%) | 234 (62.9%) |
| **Medical support staff** | 130 (68.1%) | 61(31.9%) | 28 (40.6%) | 41 (59.4%) |
| **GP** | 19 (90.5%) | * | 13 (92.9%) | * |
| **Medical Student** | 25 (47.2%) | 28 (52.8%) | 0 | 0 |

** values redacted due to the potential for identification. FY1 – foundation year 1 doctor (the first year of a two year training programme for doctors who have just left medical school); FY2 – foundation year 2 doctor (the second year of the aforementioned programme); SHO – senior house officer (a doctor in training who has completed the foundation programme and has entered a ‘core’ training programme such as core medical or core surgical training but is not yet entered speciality training); SpR – Specialist registrar (a doctor in training who has entered a speciality training programme); Consultant (a hospital doctor who has completed training); Trust grade (a doctor who is not in a training programme but is employed by the trust for provision of clinical services; may be of varying grades); Medical support staff – includes roles such as physicians associates and advanced practitioners; GP – general practitioner.*

| Variable | N vaccinated / N total (%)  11,516 / 16,433 (70.1%) | OR (95% CI) | P value | aOR (95% CI) | P value |
| --- | --- | --- | --- | --- | --- |
| **Age (years)**  ≤30  31 – 40  41 – 50  51 – 60  ≥61 | 2060 / 3490 (59.0%)  2537 / 4014 (63.2%)  2903 / 3873 (75.0%)  2989 / 3791 (78.8%)  1027 / 1265 (81.2%) | 0.48 (0.44 – 0.53)  0.57 (0.52 – 0.63)  Reference  1.25 (1.12 – 1.39)  1.44 (1.23 – 1.69) | <0.001  <0.001  <0.001  <0.001  <0.001 | 0.52 (0.47 – 0.58)  0.64 (0.58 – 0.71)  Reference  1.13 (1.01 – 1.26)  1.40 (1.19 – 1.65) | <0.001  <0.001  -  0.04  0.02 |
| **Sex**  Female  Male | 8759 / 12625 (69.4%)  2757 / 3808 (72.4%) | Reference  1.16 (1.07 – 1.25) | -  <0.001 | Reference  1.31 (1.20 – 1.45) | -  <0.001 |
| **Ethnicity**  White  South Asian  Black  Other  Not stated | 7671 / 10097 (76.0%)  2667 / 4203 (63.5%)  451 / 1034 (43.6%)  573 / 870 (65.9%)  154 / 301 (67.3%) | Reference  0.55 (0.51 – 0.59)  0.24 (0.21 – 0.28)  0.61 (0.53 – 0.71)  0.65 (0.49 – 0.86) | -  <0.001  <0.001  <0.001  0.002 | Reference  0.63 (0.58 – 0.68)  0.29 (0.25 – 0.33)  0.69 (0.59 – 0.81)  0.58 (0.43 – 0.77) | -  <0.001  <0.001  <0.001  <0.001 |
| **IMD quintile**  5 (least deprived)  4  3  2  1 (most deprived) | 3065 / 3998 (76.7%)  2580 / 3486 (74.0%)  1975 / 2850 (69.3%)  2272 / 3526 (64.4%)  1585 / 2516 (63.0%) | Reference  0.87 (0.78 – 0.96)  0.69 (0.62 – 0.77)  0.55 (0.50 – 0.61)  0.52 (0.47 – 0.58) | -  0.005  <0.001  <0.001  <0.001 | Reference  0.90 (0.81 – 1.01)  0.81 (0.72 – 0.90)  0.79 (0.71 – 0.88)  0.76 (0.67 – 0.86) | -  0.08  <0.001  <0.001  <0.001 |
| **Occupation**  Doctor  Nurse / HCA  Allied Health Professional  Admin / executive  Healthcare Scientist  Estates / Facilities  Other | 1595 / 2299 (69.4%)  4512 / 6669 (67.7%)  917 / 1281 (71.6%)  2441 / 3209 (76.1%)  608 / 806 (75.4%)  1300 / 1978 (65.7%)  143 / 191 (74.9%) | Reference  0.92 (0.83 – 1.02)  1.11 (0.96 – 1.29)  1.40 (1.24 – 1.58)  1.36 (1.13 – 1.63)  0.85 (0.74 – 0.96)  1.31 (0.94 – 1.85) | -  0.13  0.17  <0.001  0.001  0.01  0.11 | Reference  0.85 (0.76 – 0.96)  1.06 (0.90 – 1.25)  1.09 (0.95 – 1.25)  1.29 (1.06 – 1.57)  0.66 (0.57 – 0.76)  1.10 (0.77 – 1.57) | -  0.009  0.47  0.20  0.01  <0.001  0.60 |
| **Previous SARS-CoV-2 serology**  Negative  Never tested  Positive | 7306 / 10314 (75.6%)  3361 / 5611 (59.9%)  849 / 1153 (73.6%) | Reference  0.48 (0.45 – 0.52)  0.90 (0.79 – 1.04) | -  <0.001  0.15 | Reference  0.56 (0.51 – 0.60)  1.12 (0.97 – 1.29) | -  <0.001  0.14 |
| **Previous SARS-CoV-2 PCR**  Negative  Never tested  Positive | 2248 / 2886 (77.9%)  8747 / 12765 (62.3%)  521 / 782 (66.6%) | Reference  0.62 (0.56 – 0.68)  0.57 (0.48 – 0.67) | -  <0.001  <0.001 | Reference  0.70 (0.63 – 0.78)  0.71 (0.59 – 0.85) | -  <0.001  <0.001 |
| **Previous COVID-19 related work absence**  No absence  Symptomatic  Household / test and trace contact  Pregnant | 7084 / 9878 (71.7%)  2477 / 3698 (67.0%)  1931 / 2727 (70.8%)  24 / 130 (18.5%) | Reference  0.80 (0.74 – 0.87)  0.96 (0.87 – 1.05)  0.09 (0.06 – 0.14) | -  <0.001  0.35  <0.001 | Reference  0.77 (0.71 – 0.85)  0.94 (0.85 – 1.04)  0.15 (0.10 – 0.24) | <0.001  0.22  <0.001 |

**Supplementary Table 6. Univariable and multivariable analysis of factors associated with SARS-CoV-2 vaccine uptake excluding those with locum or bank contracts**
